# Clinical manifestations and mortality among hospitalized COVID-19 patients in Tanzania, 2021-2022

**DOI:** 10.1101/2023.07.13.23292643

**Authors:** Elisha Osati, Grace A. Shayo, Tumaini Nagu, Raphael Z Sangeda, Candida Moshiro, Naveeda Adams, Athumani Ramadhani, Bahati Wanja, Albert Muniko, Jeremia Seni, Mary A. Nicholaus, Kajiru G Kilonzo, Gervas Nyaisonga, Christian Mbije, John Meda, Denis Rainer, Martha Nkya, Paulo Mhame, Lucy Samwel, Liggyle Vumilia, Seif Shekalaghe, Abel Makubi

**Author notes:** Corresponding Author: Dr. Elisha Osati, (EO).

## Abstract

**Background:** There have been differential mortality rates from Corona Virus Disease of 2019 (COVID-19) in different parts of the world. It is not clear whether the clinical presentation does also differ, thus the need for this study in a Sub-Saharan African country. The aim of this study was to describe clinical manifestations and outcome of patients diagnosed with COVID-19 in selected tertiary hospitals in Tanzania.

**Methods and Findings:** A retrospective analysis of archived data from 26^th^ March, 2021 to 30^th^ September, 2022 was done for adults aged ≥18 years who were admitted in five tertiary-level hospitals in Tanzania. Information collected included socio-demographic, radiological and clinical characteristics of the patients as well as outcome of the admission (discharge vs death). Categorical variables were presented as frequencies and proportions and compared using Chi square test. Logistic regression was used to assess the relationship between COVID-19 mortality and the collected variables. Out of 1387 COVID-19 patients, approximately 52% were males. The median age was 60 years [(IQR)= (19-102)). The most common symptoms were dyspnea (943,68%), cough (889, 64%), fever (597,43%) and fatigue (570, 41%). In hospital mortality was (476, 34%). Mortality significantly increased with increasing age, being the most in age >90 years [aOR (95% CI) =6.72 (1.94-20.81), P<0.001. Other predictors of death were not possessing a health insurance, [aOR (95% CI) = 2.78 (2.09-3.70), P<0. 001], dyspnea [aOR (95% CI) = 1.40(1.02-2.06), P=0.03]; chest pain, [aOR (95% CI) = 1.78 (1.12-3.21), P=0.03]; HIV positivity, [aOR (95% CI) = 4.62 (2.51-8.73), P<0.001]; neutrophilia, [aOR (95% CI) = 1.02 (1.01 – 1.03), P=0.02]; none use of ivermectin, [aOR (95% CI) = 1.46 (1.09 – 2.22), P=0.02] and non-use of steroid, [aOR (95% CI) = 1.40 (1.2 – 2.5), P=0.04]. Retrospective nature of this study which based on documented patients records, with a large number of patients left out of the analysis due to missed data, this might in a way affect the results of the present study.

**Conclusions:** The most common presenting symptoms were dyspnea, cough and fever, just as what was common elsewhere in the world. Mortality increased significantly with age, in HIV-infected patients, in those without a health insurance, those presenting with dyspnea, chest pain, or neutrophilia and those who did not use steroid or ivermectin. Clinicians should actively look for the predictors of mortality and take appropriate management to reduce mortality.

## Introduction

By March 2023 a total of 760 million infections from SARS-COV 2 and about 6.9 million deaths had been recorded globally ^1^, making Corona virus disease of 2019 (COVID 19) the number one killer from a single infectious cause, surpassing tuberculosis^2^.

Compared to the Americas, Europe and Asia, African countries including Tanzania have recorded low incidence of symptomatic COVID-19 infection as well as mortality due to yet unexplained reasons. Nevertheless, the COVID-19 pandemic has added to the disease burden to Sub-Saharan Africa (SSA), which also has the highest numbers of other infectious diseases and escalating rates of non-communicable diseases^3^.

COVID-19 illness may range from a period of no symptoms to severe COVID-19 pneumonia, with acute respiratory distress syndrome (ARDS) possibly leading to death.

Although COVID-19 is primarily a respiratory disease, recent data suggest that it can lead to cardiovascular,^4–7^ hematological,^6, 8^ hepatic,^6^ neurological,^7, 9^ renal,^6, 10^ and other complications. ^6^ Fever and cough are the most common presentations, seen in 60-87% and 72-85% of the patients with COVID-19 respectively. ^7, 11–13^ Fatigue has been reported in 20-36% of the cases.^14, 15^ Dyspnea is the most frightening symptom and is seen in about a quarter of the patients. ^14, 15^ Neurological symptoms include headache in about 34% of the patients ^7, 16, 17^ while about 7% of the patients had reported impaired sense of smell and taste. ^13, 18^ Other symptoms reported in acute COVID-19 included muscle pain in up to 34% of the patients, nausea (4.1%,), anorexia (2.6%,) and sore throat (1.6%). ^11, 13, 14^ Other reported symptoms include, but are not limited to diarrhea, vomiting, abdominal pain rhinorrhea and dizziness. ^7, 19^ Hypoxemia has been strongly associated with worsening of clinical outcomes. Xien J, *et al* in a study done in 2020 in Wuhan China found that oxygen saturation (SpO_2)_ of less than 90.5% at admission was related to an almost 3-fold increased risk of dying^20^.

The pulmonary abnormalities seen in the chest imaging have been commonly bilateral peripheral ground-glass opacities (GGO). Consolidation developed later in the course of COVID-19 illness.^21^ An earlier imaging study in Wuhan, China reported that bilateral lung abnormalities were the predominant findings in 79% of patients followed by peripheral abnormalities (54%)Click or tap here to enter text. GGO was found in 65% of the patients and mainly involved the right lower lobes (27%). ^21^

Leukopenia and lymphopenia are the most common laboratory findings found in patients with COVID-19.^6^ though they are non-specific. Other abnormal laboratory findings include elevated levels of lactate dehydrogenase, C-reactive protein, aminotransferase, D-dimers and ferritin.^6, 7^ Patients with elevated levels of D-dimer were found to have a 3-fold risk of mortality among COVID-19 patients in a study done in Chennai India. ^7, 22^

High cytokine levels (IL-2R, IL-6, IL-10, and TNF-α), and high lactate dehydrogenase (LDL) level were significantly associated with severe COVID-19 on admission in a retrospective study done in two hospitals in Wuhan, China. ^23^

Several factors have been associated with adverse outcomes and mortality among COVID-19 patients. These include male sex, age of more than 55 years, ^24, 25^ pre-existing comorbidities, ^24, 26,27^ hypoxic state at admission, radiological abnormalities, ^28^ abnormal laboratory results,^29^ and bio-markers of multiple organ failures.^30^ The presence of these factors has been used by physicians to predict the severity of COVID-19 and the risk of death. ^29^

A study done in Iran among 205,654 COVID-19 patients reported that patients who were 60 years or older or were male or presented with low SpO_2_ levels at the time of admission were more likely to die of COVID-19.^31^ Lower socio-economic status was also associated with increased risk of death from COVID-19.^31, 32^ Some studies conducted early during the pandemic reported more severe COVID-19 for smokers. ^33, 34^ However one study reported that smoking was not associated with lower oxygen saturation or death in COVID-19. ^35^ Of note, these studies did not consider the important confounding factors like age, sex, and pre-existing comorbidities.^33, 34^

Comorbidities such as obesity, diabetes, and hypertension have been reported to increase the risk of developing more severe COVID-19. ^7,36^ A retrospective multi-center study in Pakistan among patients with severe COVID-19 showed that 19% of these cases were associated with one or more comorbidities.^14^ The most common comorbidities were hypertension (affecting 7.7% of the COVID-19 patients), diabetes mellitus (4.6%), cardiovascular diseases (2.6%), asthma (1.6%), and other co-morbidities (2.6%). ^14^ The CDC has included sickle cell disease, asthma, and pregnancy as risk factors for severe COVID-19^37^.

In the USA fatality rates among COVID-19 patients were found to be higher in cardiovascular conditions (10.5%) followed by diabetes mellitus (7.3%), COPD (6.3%), hypertension (6.0%), and cancer (5.6%),^35^ as compared with fatality rate of less than 1% in COVID-19 without pre existing comorbidities^35^.

Currently treatment of SARS-CoV2 infection relies mostly on symptomatic treatment and supportive care of the presenting problems.^18^ Management strategies have been directed to address inflammation, hypercoagulability, oxygenation, vitamin and supplements, restoration and maintenance of hydration, prophylactic antibiotics and promising antivirals.^18^ Administration of systemic steroids in patients with severe COVID-19 has been shown to reduce the risk of mortality by 64%.^38^ Remdesivir has been shown to lower the risk of mortality, accelerate patients’ recovery, and reduce progression to invasive ventilation, compared to best supportive care among hospitalized COVID-19 patients requiring any or low supplemental oxygen at baseline.^39^ Invasive ventilation has been associated with 36% of mortality in the ICU among severe COVID-19 patients.^25^ Ivermectin has been reported by Caly, *et al* to inhibit SARS-CoV2 in vitro and has been used during acute COVID-19. ^40^

COVID-19 vaccines reduce the severity and transmissibility of SARS-CoV2 infection. ^41^ It has been reported in the WHO Coronavirus Dashboard report with Vaccination of September, 2022 that about 20% of vaccinated individuals in the general population had an infection after successful vaccination^17^.

Our knowledge of clinical description, risk factors and outcomes of COVID-19 in Tanzania is limited to reports from other countries. This study therefore aimed to describe clinical manifestations and outcomes of patients diagnosed and hospitalized with SARS-COV2 in a Tanzanian population.

## Materials and methods

### Design and setting of the study

This was a retrospective analysis of archived data of COVID-19 patient in five tertiary hospitals in Tanzania, namely Muhimbili National Hospital (MNH) Upanga and Mloganzila campuses in Dar es Salaam city representing the coastal zone, Kilimanjaro Christian Medical Center (KCMC) in Kilimanjaro representing the northern zone, Bugando Medical Center (BMC) in Mwanza representing the lake zone, Benjamin Mkapa Hospital (BMH) in Dodoma representing the central zone and Mbeya Zonal Referral Hospital (MZRH) in Mbeya representing the southern highlands zone. The hospitals were selected conveniently due to their capacity to accommodate and manage severe respiratory diseases including COVID-19.

We studied archived data of patients aged 18 years or older who were admitted in the participating hospitals from 26^th^ March, 2021 to 30^th^ September, 2022 with COVID-19 confirmed by Polymerase Chain Reaction (PCR) test. PCR tests/reagents were not always available in the country before March, 2021 thus, patients seen in those hospitals during that time were not part of this study. Patients who have incomplete data of more than 10% were excluded from the analysis. Approval for this study was obtained from the Muhimbili University of Health and Allied Sciences (MUHAS) institution review board, with approval number (MUHAS-REC-10-2022-1404) and since this was retrospective study an exemption was obtained and no consent was required.

### Data collection procedures

Dat was collected for the period of six months from November, 2022 May 2023. We searched patients’ data from Hospitals’ record departments to identify patients who were hospitalized with the diagnosis of either confirmed or suspected COVID-19 disease or other diagnoses which in our experience were often used instead of COVID-19. We obtained file numbers of these patients from the records departments and used them to search for COVID-19 PCR results from the hospitals’ records and/or from the National Public Health Laboratory (NPHL). Only data of patients with positive PCR tests were considered for this study. File numbers were also used to obtain both hard copy files and electronic data of the patients. Electronic Clinical Research Forms (CRF) created in Research Electronic Data Capture (REDcap) software were used to document patients’ data from both hard copy files and electronic databases. Patients’ names and registration numbers were not entered in a database. Patients were assigned a special study number which were entered in the database. The database was only accessed by the study team with special passwords. The information collected included sociodemographic data and vital signs (respiratory rate, oxygen saturation and heart rate), symptoms (respiratory, cardiovascular, gastrointestinal, and central nervous system), duration of hospital admission, comorbidities, laboratory characteristics including CD4 and viral load, treatment modalities, and mortality.

### Statistical analysis

With our sample size of 1387 there is 80% power to detect a risk factor with prevalence 0.06 if it has an odds ratio of at least 1.4. Study data were collected and managed using REDcap software hosted at MUHAS ^42^ Analysis was done using STATA version 17. Socio demographic, clinical and radiological characteristics of the patients were categorized, presented as frequencies and proportions and compared using Chi square test or Fishers exact test. We used Fishers exact test for variables with less than 25 responses, i.e., variables with low diversity (e.g., binary variables with a prevalence of under 2% of one category). Continuous variables were presented as median and interquartile range and compared by Wilcoxon rank sum test. Logistic regression was used to assess the relationship between sociodemographic factors, clinical presentations, comorbidities; and treatment modalities and COVID-19 outcomes. Covariates for the multivariate logistic regression were selected using a P-value threshold of 0.05 from the univariate analysis. Some potential risk factors that have been widely reported in the literature were forced into the multivariate model. For example, sex had a p-value greater than 0.05, it was entered into the multivariate model due to its clinical significance. Variables with low diversity (e.g., binary variables with a prevalence of under 2% of one category) and variables with large numbers of missing data were not included in the model, regardless of their meeting other criteria. P-value < 0.05 was considered significant in all analyses.

## Results

A total of 10,237 suspected COVID-19 patients were admitted in the participating hospitals from 26^th^ March 2021 to 30^th^ July 2022. A total of (6,875, 67%) PCR results were not reported and (1,206, 12%) had negative PCR results. Only (2,156, 21%) were confirmed to have SARS-CoV-2 infection by PCR, of whom (1387, 64%) had complete data and were included in the final analysis (Fig 1).

**Figure 1:**
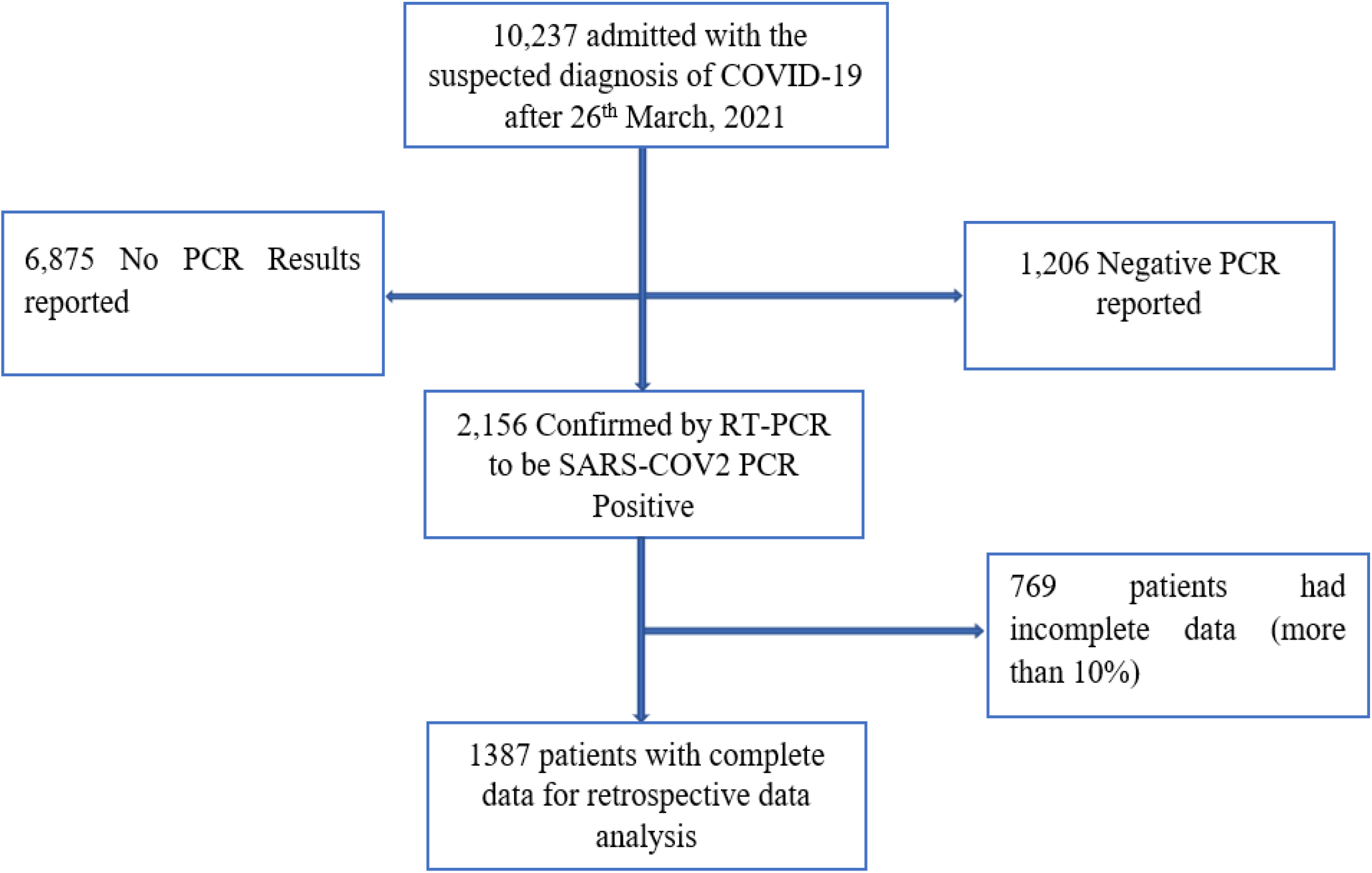
Patients screening and recruitment flow.

Out of 1387 patients included in the analysis 48% (669/1387) were from Muhimbili National Hospital (MNH) Dar es Salaam, 23% (313/1387), from Kilimanjaro Christian Medical Center (KCMC) in Moshi Kilimanjaro, 15% (209/1387) from Mbeya Zonal referral Hospital (MZRH) in Mbeya, 8% (111/1387) from Bugando Medical Center (BMC) in Mwanza region, and 6% (85/1387) from Benjamin Mkapa Hospital (BMH) in Dodoma (Fig 2).

**Figure 2:**
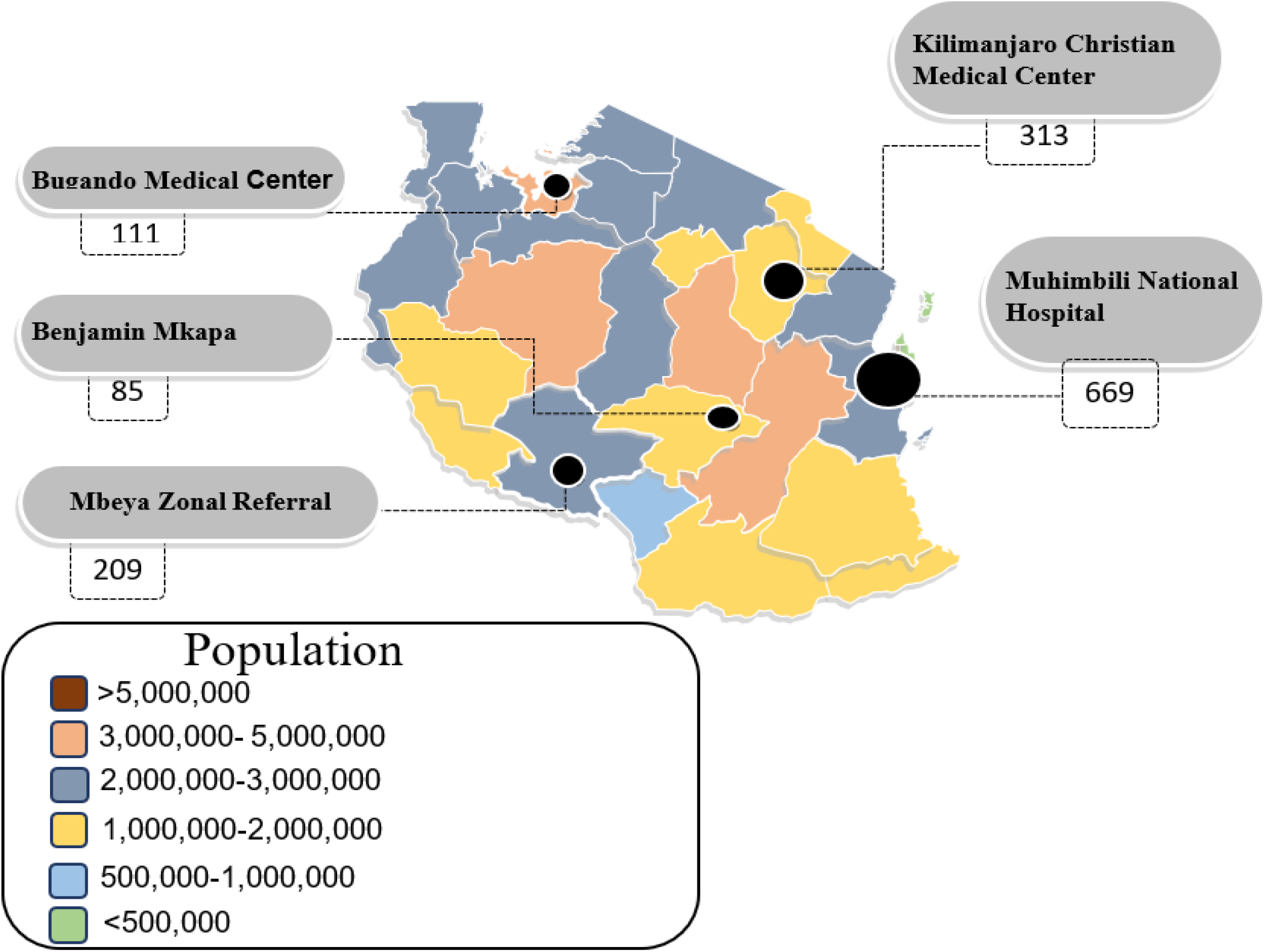
Number of studied patients in each participating hospital and estimated population in the regions of domicile.

## Characteristics of the patients at admission to the hospital

### Sociodemographic

The median age of the patients was 60 (19,102) years with (501, 36%) in the age group 60-74 years. More than a half were males (722, 52%). Most of the patients had finished either primary (36, 26%) or secondary school (372, 27%). Nearly half (626, 45%) were covered by a health insurance. (Table 1). In-hospital mortality was (476, 34%). (Table 1).

**Table 1:**
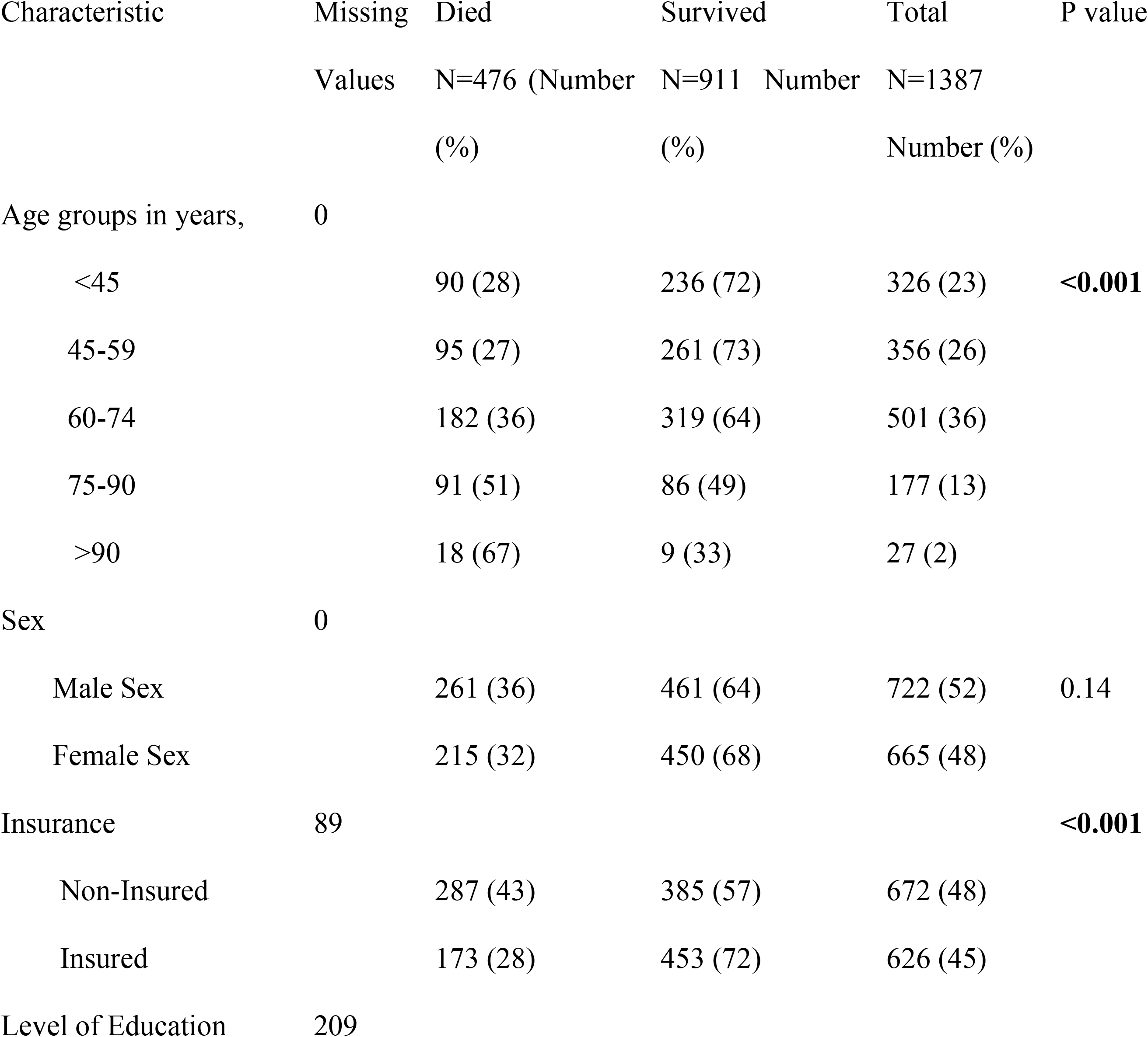
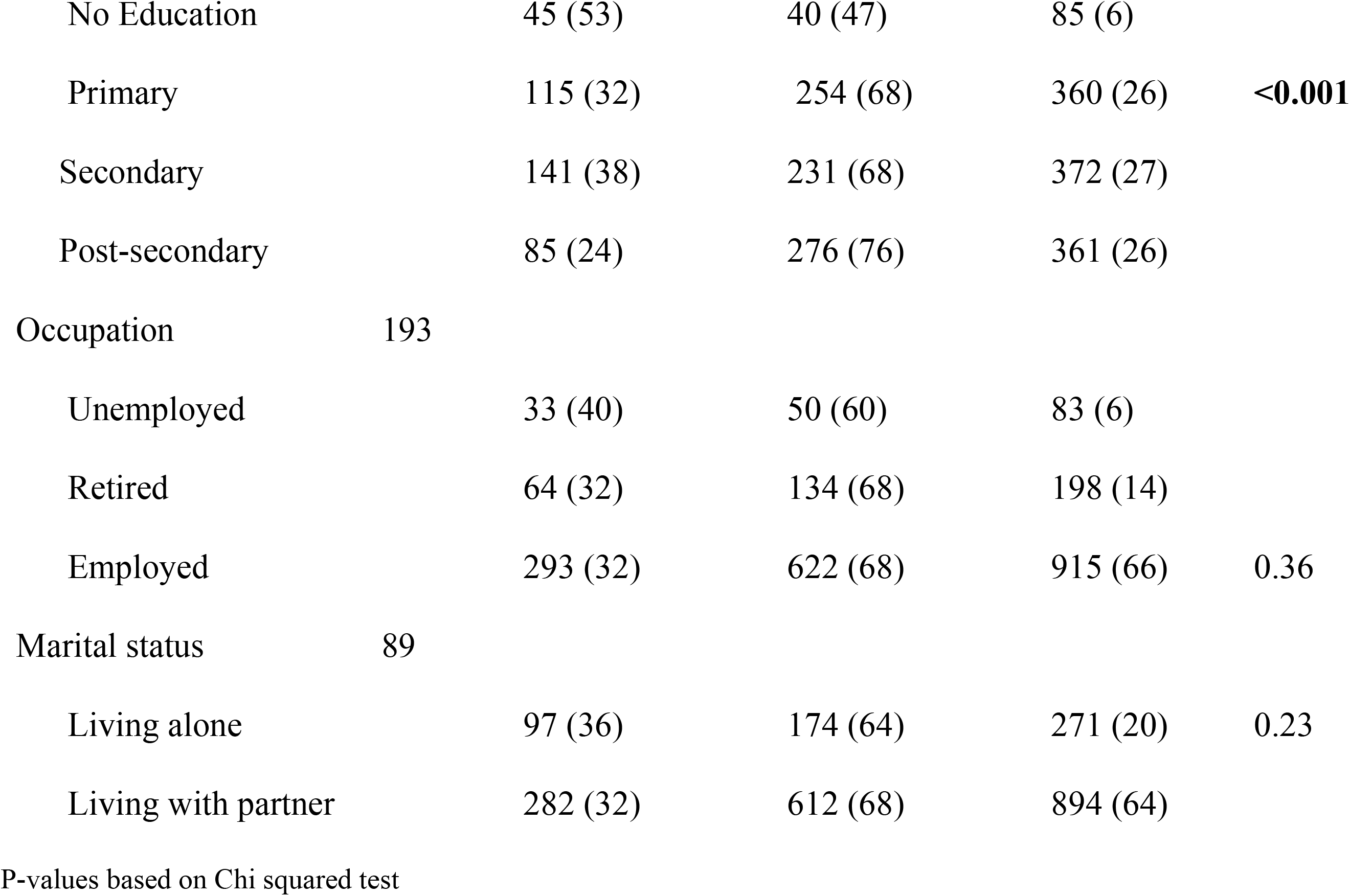
Sociodemographic characteristics at admission of patients hospitalized with COVID-19 at admission in Tanzania, 2021-2022, N=1387.

### Comorbidities, symptoms, laboratory, and radiologic findings at hospital admission

The most common comorbidities among the 1387 patients were hypertension (238, 40%), and diabetes mellitus (DM) (127, 40%). The most common symptoms were dyspnea (943, 68%), cough (889, 64%), fever (597, 43%), fatigue (570, 41%), chest pain (364, 26%), and headache (252, 18%). More than half of the patients had oxygen saturation (SpO_2)_ of at least 95% (777, 56%). The number with tachypnea was (138, 10%), and (683, 49%) had tachycardia. Almost a half of the admitted patients (683, 49%) had lung infiltrates as reported in chest x-rays (Table 2).

**Table 2:**
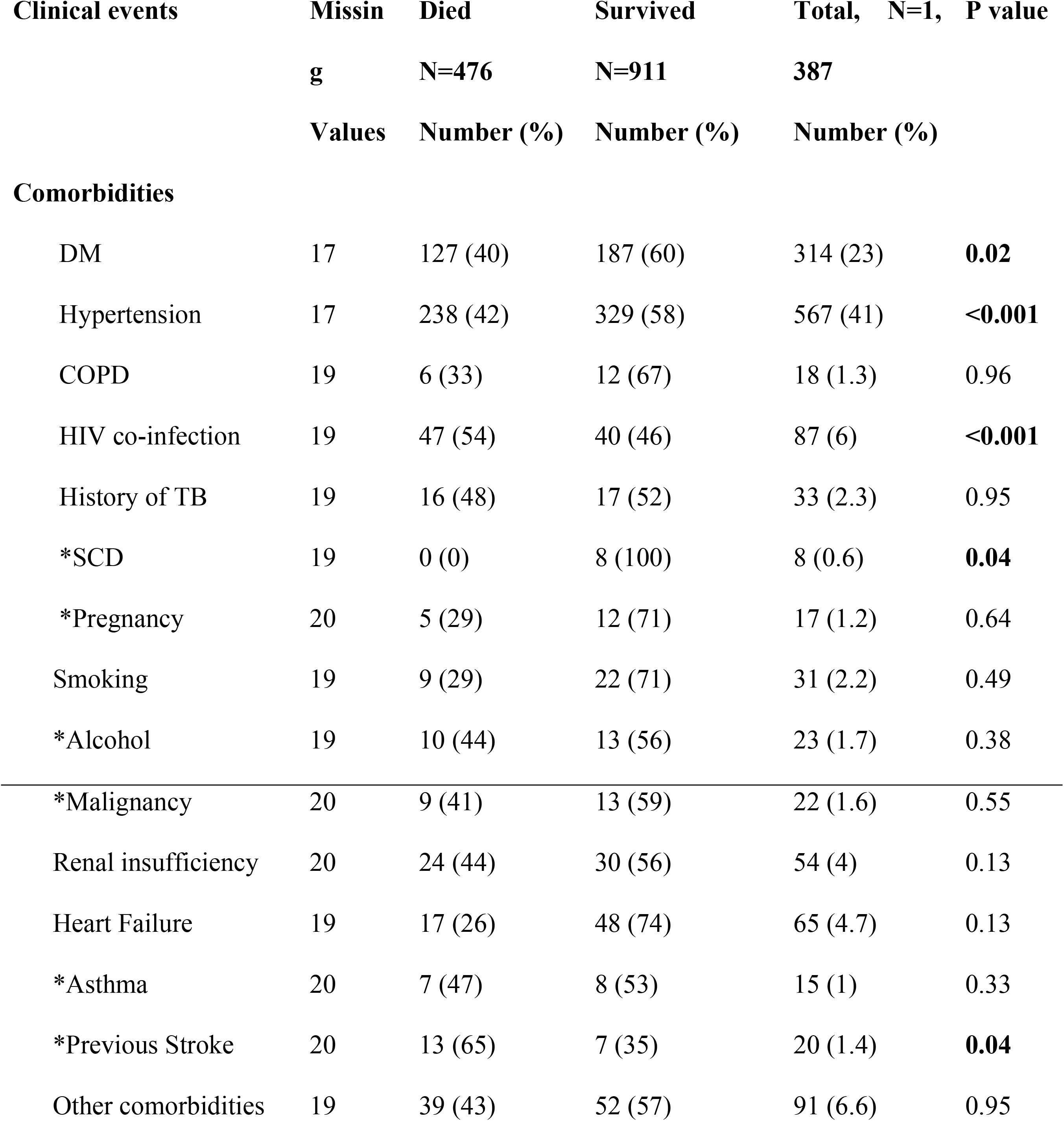
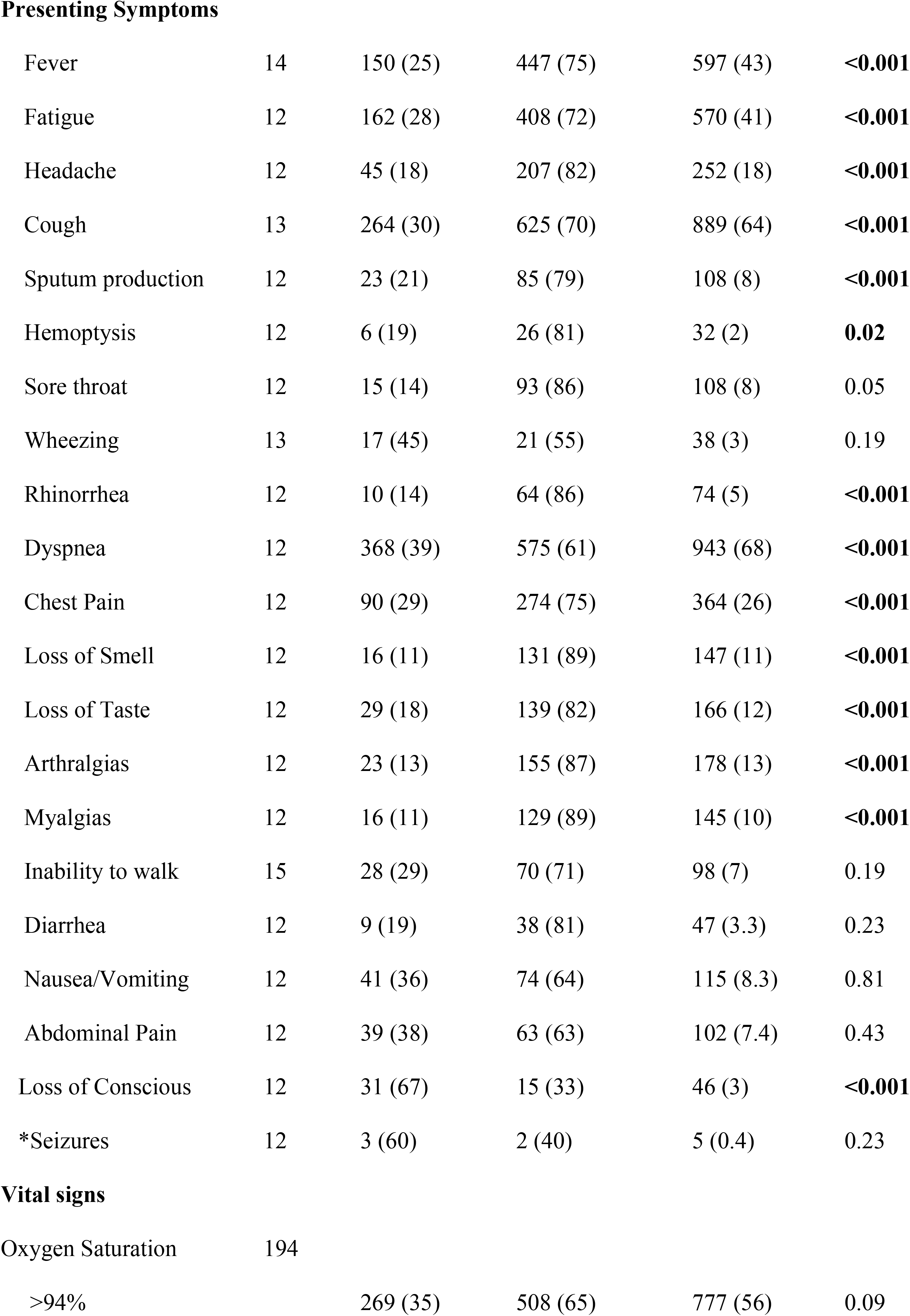
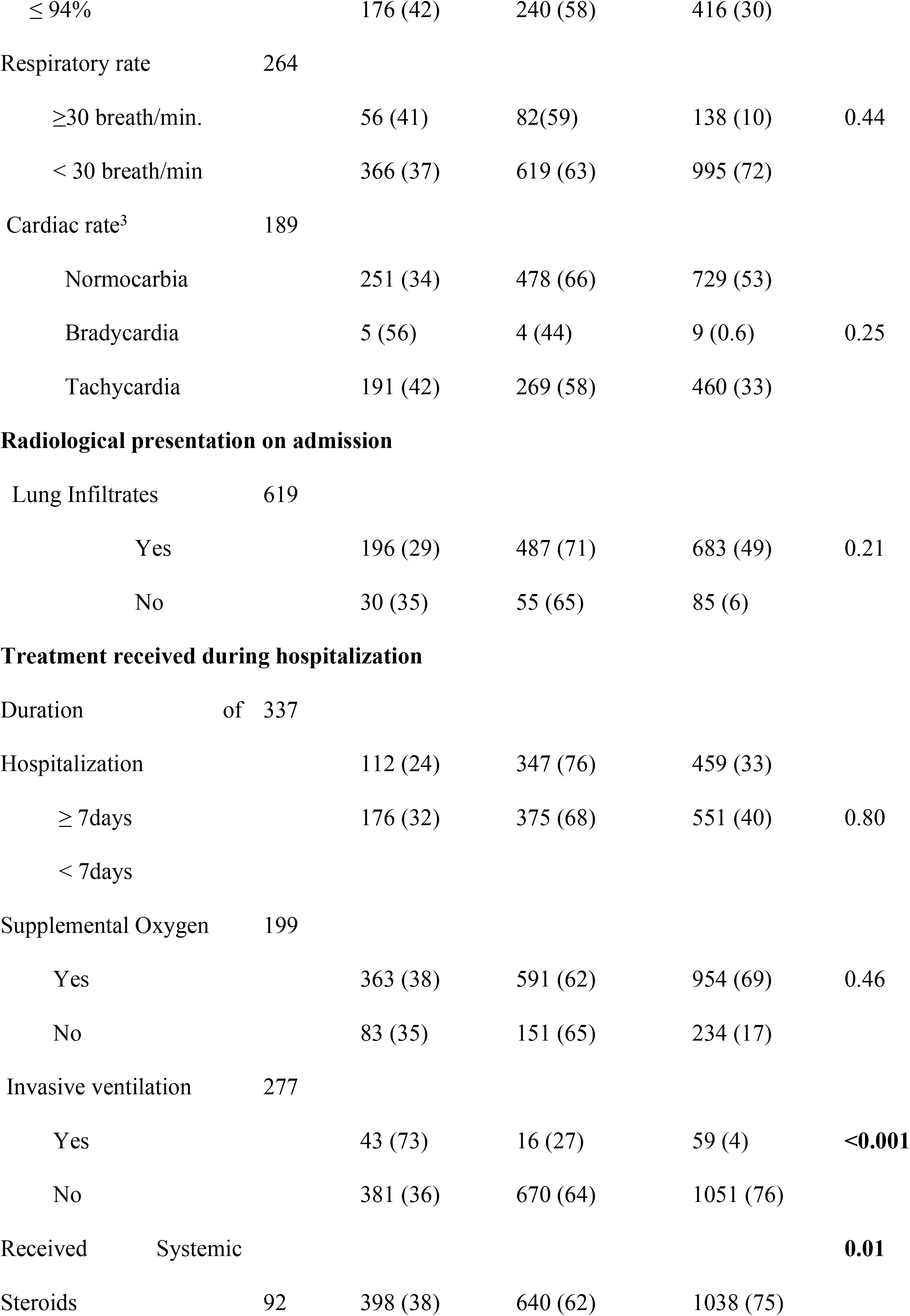
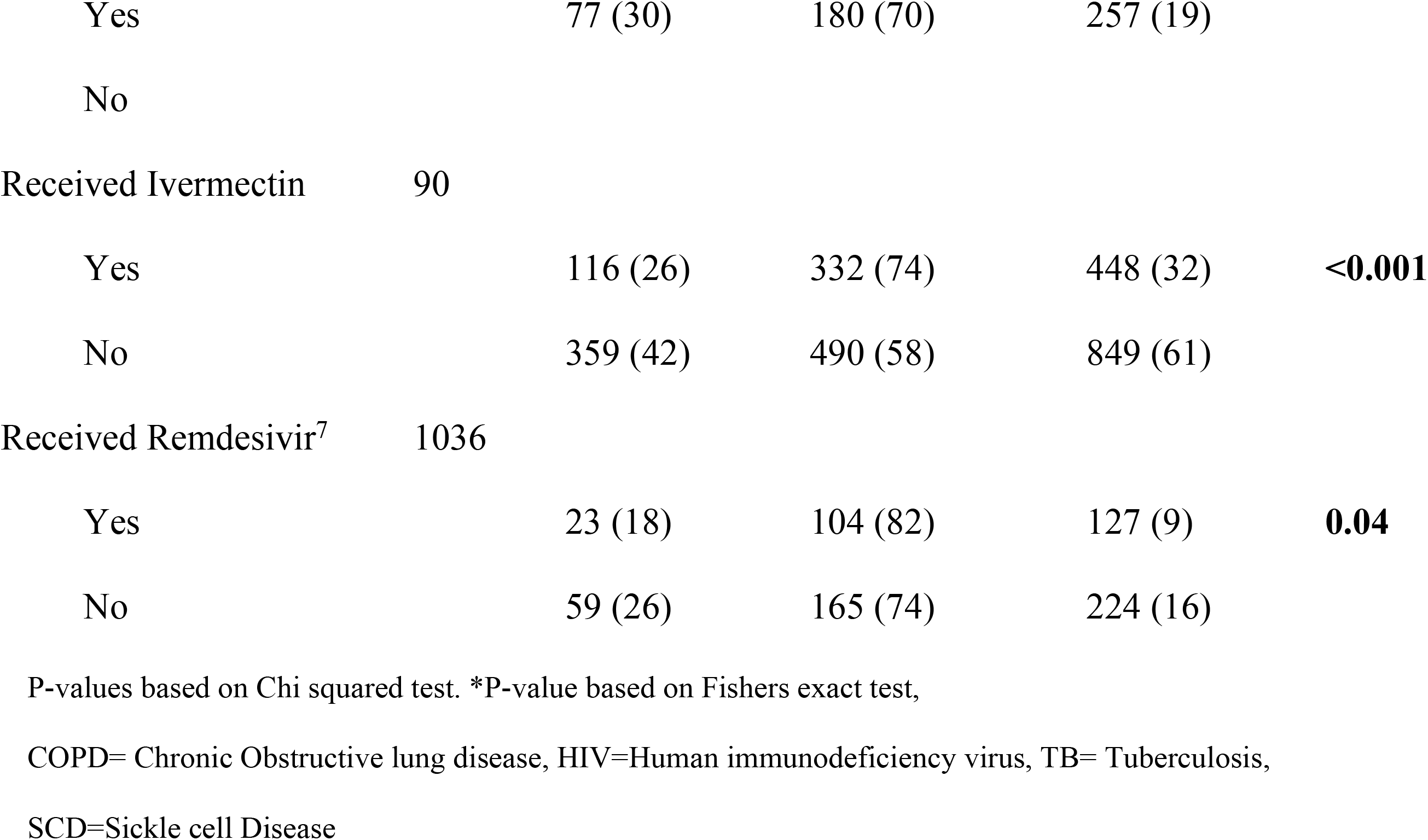
Proportions of clinical presentations, comorbidities and treatment modalities of patients hospitalized with COVID-19 at admission, Tanzania, 2021-2022, N=1387.

### Hospital treatments received

Although most of the patients had normal SpO^2^ at admission, (954, 69%) received supplemental oxygen during their hospitalization. Three-fourths received steroids (1038, 75%), (448, 32%) received ivermectin and (127, 9%) received remdesivir. Only (59, 4%) received invasive ventilation.

There were many observations with missing data on the laboratory investigations as shown in Table 3 below. However, patients who died compared to patients who survived had higher median values of CRP, [Median (IQR)=54.7 (14.8, 124) mg/L, P=0.03]; D-dimer, [Median (IQR)= 4.4 (1.1, 148.5) μ/mL, P=0.02]; white blood cell count (WBC), [Median (IQR)= 9.6 (6.8, 13.7) 109/L, P <0.001]; absolute neutrophil count (ANC), [Median (IQR)=7.8 (4.9, 11.2)109/L, P <0.001]; fasting blood glucose (FBG), [Median (IQR)= 8.3 (6.2, 13.9) mmol/L, P<0.001]; Creatinine, [Median (IQR)= 118 (87, 177) μmol/L, P<0.001]; and Blood Urea Nitrogen (BUN), [Median (IQR)= 7.8 (5.6, 12.2) mmol/L, P<0.001] Table. 3

**Table 3:**
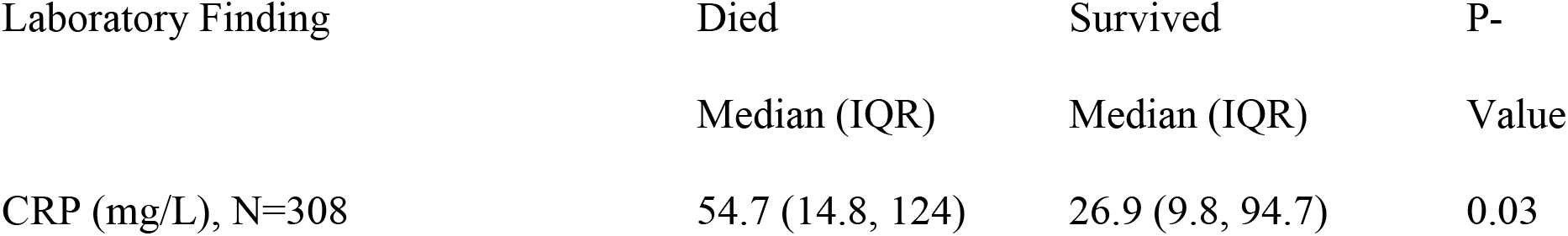
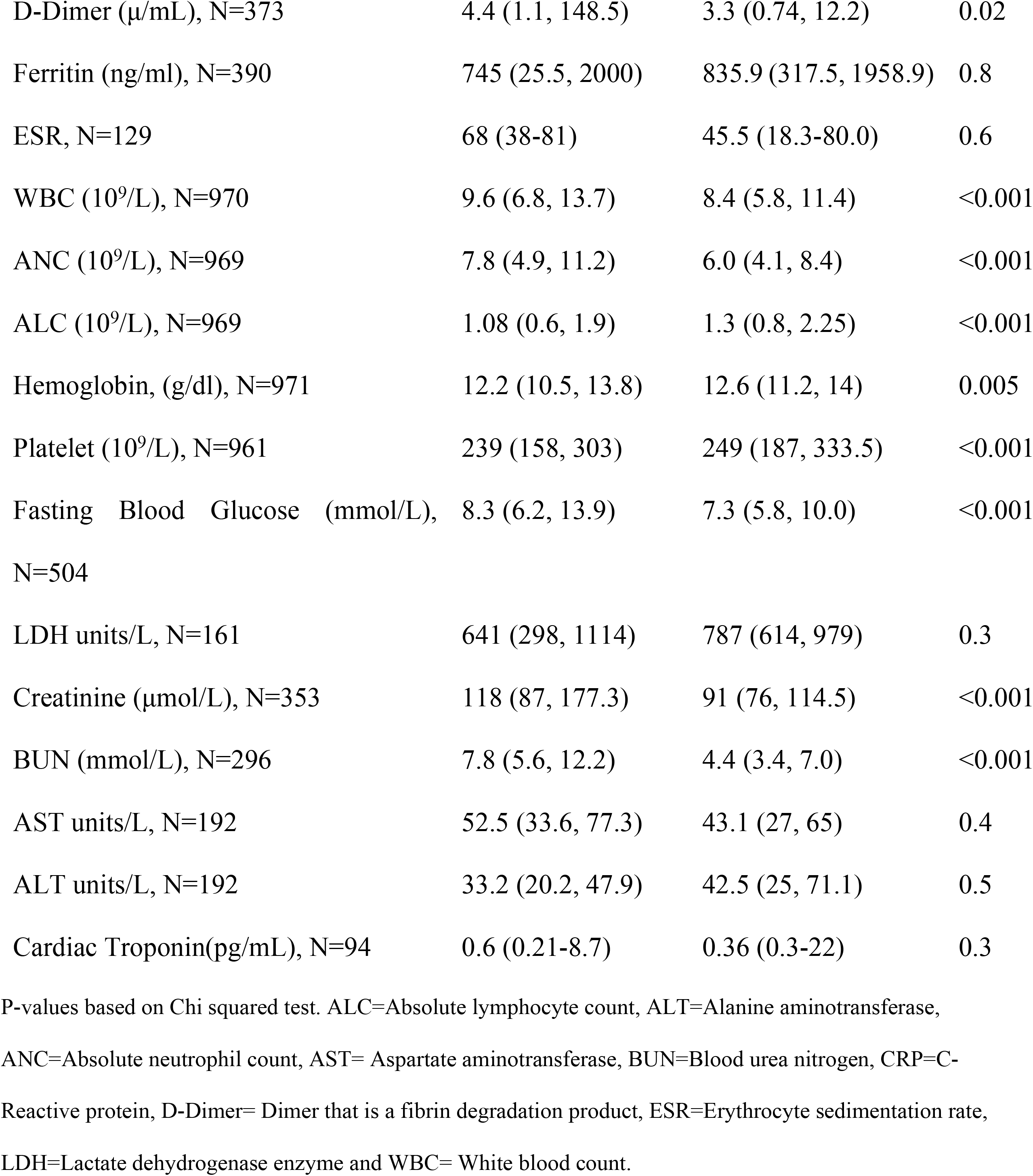
Laboratory investigations of patients hospitalized with COVID-19 at admission, Tanzania, 2021-2022.

### Results of multivariate models

In univariate analysis, mortality was associated with increasing age (with odds ratios increasing significantly with increasing age), lack of health insurance, HIV infection, dyspnea, chest pain, higher neutrophilia, none use of steroid and none use of ivermectin, all p value < 0.05 (Table 4). After controlling for other factors in multivariate analysis, compared to age group of less than 45, the odds of dying were almost 3 folds in age group 60-74 years, [aOR (95% CI) =2.72 (1.05-6.71), P<0.001], four folds in age group 75-90 years, [aOR (95% CI) =3.80 (1.31-10.80), P<0.001] and 7 folds in elders above 90 years, [aOR (95% CI) =6.73 (1.92-20.81), P<0.001]. The odds of death were almost 3-folds higher among uninsured patients compared to those with health insurance, [aOR (95% CI) = 2.78 (2.09-3.70), P<0. 001]. The odds of death were 40% higher in patients with dyspnea than patients without dyspnea [aOR (95% CI) = 1.40 (1.02-2.06), P=0.03]. The odds of death were 78% higher in patients with chest pain than patients without chest pain, [aOR (95% CI) = 1.78 (1.12-3.21), P=0.03]. The odds of death were over 4-folds higher among patients with HIV co-infection compared to HIV negative, [aOR (95% CI) = 4.63 (2.52-8.74), P<0.001]. Higher level of neutrophils had 2% increase risk of death compared patients with low levels of neutrophils, [aOR (95% CI) = 1.02 (1.01 – 1.03), P=0.02]. None use of ivermectin was associated with 58% mortality, [aOR (95% CI) = 1.46 (1.09 – 2.22), P=0.02]. None use of steroid was associated with 60% mortality, [aOR (95% CI) = 1.40 (1.27 – 2.53), P=0.04] (Table 4).

**Table 4:**
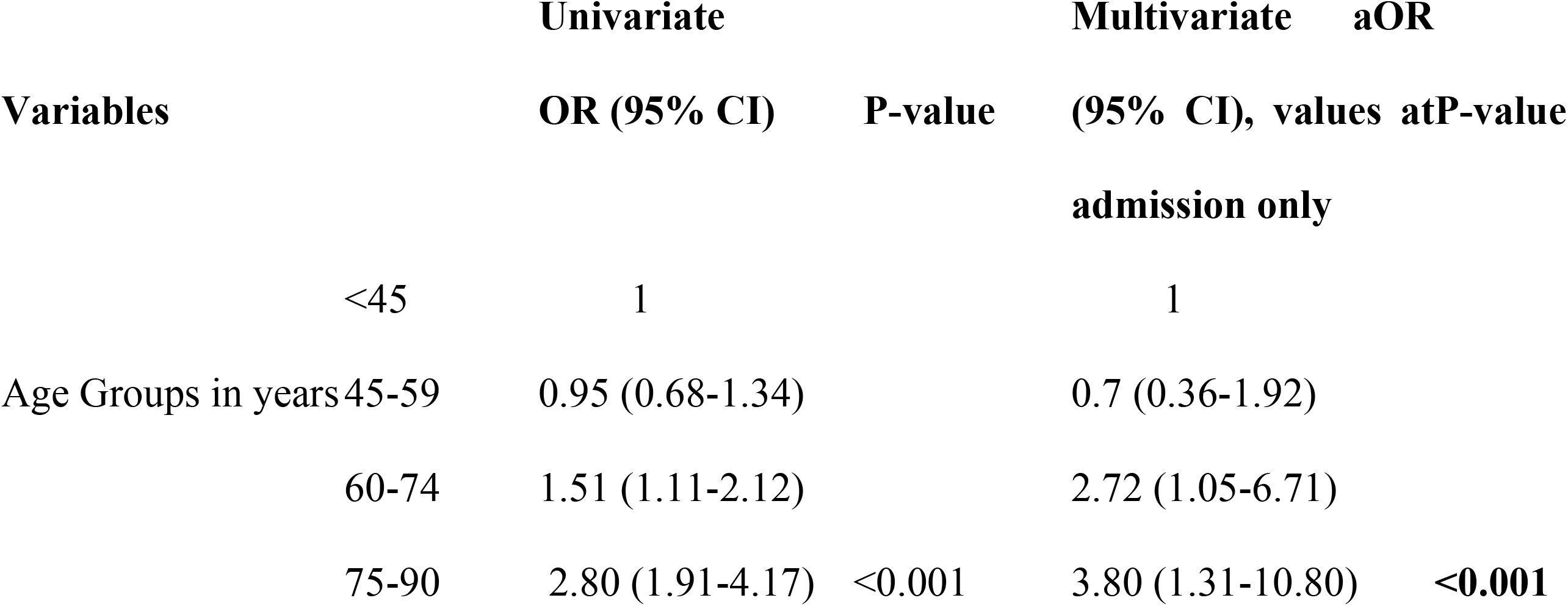
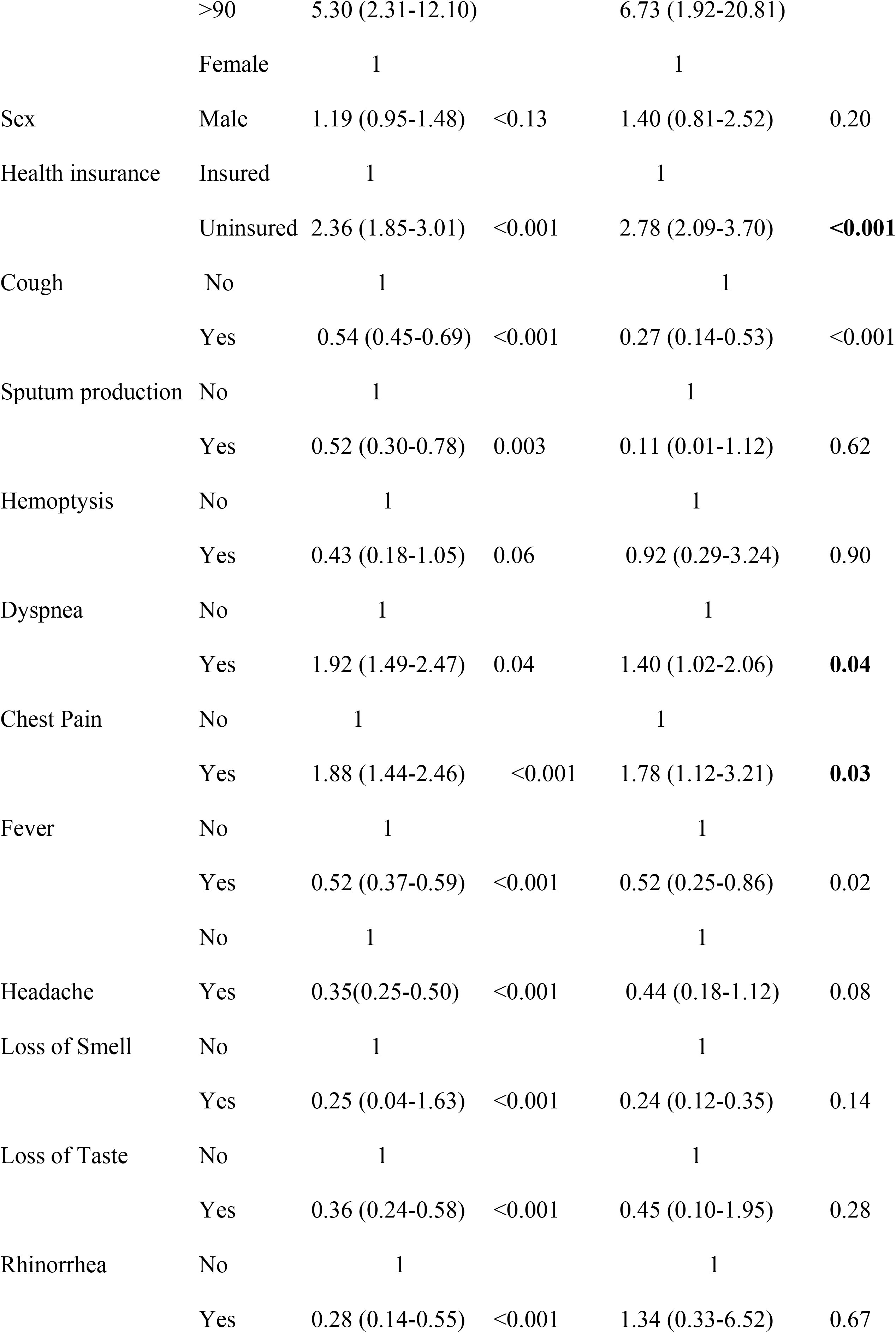
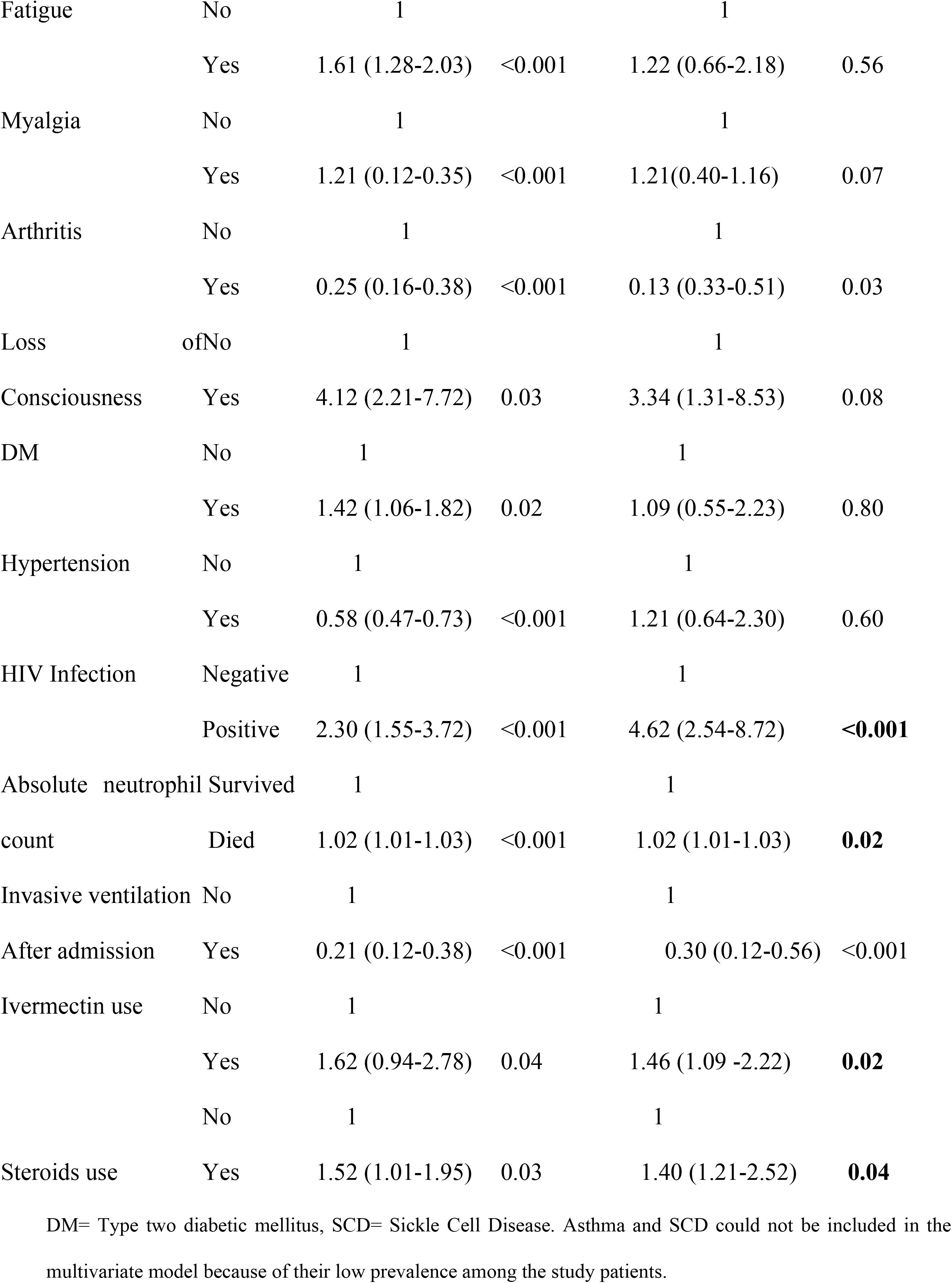
Sociodemographic, clinical and treatment predictors for mortality, among patients hospitalized with COVID-19 at admission, Tanzania, 2021-2022, N=1387.

## Discussion

We report on clinical manifestations and outcome among 1387 patients admitted with confirmed COVID-19 to five hospitals in Tanzania. Almost half the study participants were from Muhimbili National Hospital (MNH) in the largest metropolitan city, Dar es Salaam, followed by Kilimanjaro Christian Medical Center (KCMC) in Moshi Kilimanjaro representing Northen part of Tanzania, Mbeya Zonal referral Hospital (MZRH) in Mbeya city representing the Southern Highland part of Tanzania. Bugando Medical Center (BMC) in Mwanza city at the lake zone and Benjamin Mkapa Hospital (BMH) in Dodoma, the central zone of Tanzania each having less than ten percent of participants recruited in the study. Few numbers of patients from BMH Dodoma could be due to low population density as compared for example with MNH which is in Dar es Salaam (5,383,728 population). Again, there were significant missing data from hospitals like BMC and BMH.

Similar to other studies ^7, 24, 25^, the majority of patients in this study were male. One-third of COVID-19 patients died in the hospital. The odds ratio of death increased significantly as patients age increased above 60 years. There were almost 3-folds increased odds of dying in the age group 60-74 years while the odds of dying were almost 7-folds in the age above 90 years. The relationship between age and COVID-19 deaths in this study reflects not only WHO data,^1^ but also the findings of other studies of COVID-19 done elsewhere. For example, a study done in Sudan by Hasabo E, *et al,* ^7^ Lombardy Region, Italy by Cecconi M, *et al* ^24^ and Grasselli G, *et al* ^25^ which indicated that majority of confirmed COVID-19 deaths were among patients aged above 60 years.

Male sex has been found to be an independent factor associated with severe COVID-19 and mortality.^14, 15, 23^ In our study sex was not found to be a risk factor for death.

Patients who died were more likely to be unemployed, uninsured and with no formal education. The risk of deaths was almost 3-fold in uninsured group compared to insured one. Being unemployed, no formal education and being uninsured are proxy indicators of poor socioeconomic status which have been reported to be one of the risk factors of adverse outcomes and mortality from COVID-19.^31, 32^

Obesity, diabetes, and hypertension have been shown to increase the risk of developing more severe COVID-19 and mortality.^43^ Data on obesity among COVID-19 patients were not recorded consistently in patients’ file in our study. The leading comorbidities were hypertension followed by DM, though they were not found to be predictors of mortality in this study.

COVID-19 co-infected with HIV had almost 5-folds of death compared to HIV negative. This finding seems to be higher than what was reported in a review published in The *Lancent* in May 2022 which showed 38% greater odds of in-hospital death for HIV-infected patients compared to HIV negative COVID-19 patients.^44^Again, the findings in the present study have shown higher odds of death than the findings of a meta-analysis on the outcome of patients with COVID-19 and HIV co-infected individuals which showed that individuals with HIV had increased chance of hospitalization for COVID-19 with 2-folds increased odds of death regardless of CD4+ and HIV Viral load^45^.

Almost two-third of admitted COVID-19 patients in this study presented with dyspnea and cough which is similar to studies done elsewhere.^20^ In contrast to the findings of our study which showed two-thirds of COVID-19 patients to have dyspnea, other studies found only a quarter of the hospitalized patients had dyspnea.^14, 15^ Perhaps because patients don’t come to hospital till, they have severe symptoms. Other symptoms in order of importance were fever and fatigue which were present in almost a half of patients, chest pain was present in a quarter of COVID-19 patients.

Fever was preset in a half of patients in our study, this was low compared to findings of studies done in Saudi Arabia, USA, and China.^11–13, 46^ For example, a study of 370,000 confirmed COVID-19 patients in United States, reported that fever was present in 70% of patients.^46^ Fatigue was found in almost a half of our study patients, this is a bit higher compared to reports in other studies which showed fatigue to be in one-third of COVID-19 cases.^14, 15^. This could be due to the fact that patients came to the hospital late and in a severe form due to the denial of existence of COVID-19 by authorities in Tanzania during the 2^nd^ phase of the pandemic.

Significantly, more deaths were observed in patients with dyspnea and chest pain. Dyspnea was found to have 40% higher risk of death in the current study. This is the same ass the findings of a study done in Sudan by Hasabo E, *et al* in 2021, which found dyspnea to be associated with death by a factor of two in multivariable analysis^7^ Chest pain had 78% higher risk of death in our study and has been strongly associated with worsening of clinical outcomes in other studies.^7, 20^ Dyspnea and chest pain in COVID-19 patients occurs because of the virus induced inflammatory responses leading to lung damage; The damage is evidenced by acute respiratory distress syndrome (ARDS) with diffuse alveolar damage (DAD), diffuse thrombotic alveolar microvascular occlusion, and inflammatory mediator-associated airway inflammation.^47, 48^ The combination of these three pathogeneses impairs alveolar oxygenation, lead to hypoxemia, and respiratory acidosis. If these hypoxic states are not treated may result into death from respiratory failure, or sequelae of permanent lung damage. ^47–49^

In the present study people who died had significantly higher median CRP, D-dimer, white blood cell count (WBC), absolute neutrophil count (ANC), higher fasting blood glucose (FBG), serum creatinine, blood urea nitrogen (BUN) compared to survivors.

Among the laboratory findings only ANC was found to be a predictor of mortality by at least 2%. This is in harmony with the findings of two studies from Wuhan China done by Yang X, *et al* ^6^ and Li X *et al* ^23^ which reported that adverse outcome of COVID-19 was associated with neutrophilia.^8^ Again higher absolute value of neutrophils could mean bacterial superinfection in COVID 19 patients, which perhaps increased the chance of mortality.

A half of COVID-19 patients in our study presented with lung infiltrates as reported by chest X-rays. Radiological documentations in this study did not take into account distribution of the opacities, however, more deaths were observed among patients without opacification on chest X-ray without statistical significance. More than 50 % of the patients’ files in this study had no reported radiology findings.

Regarding treatment modalities that were given to admitted COVID-19 patients, three-quarters of patients received systemic steroids and supplemental oxygen. Ivermectin was used by one-third of patients and only one-tenth used Remdesivir.

The odds of death were found to be significantly lower in patients who used ivermectin. Caly, *et al* found that ivermectin can inhibit SARS-CoV2 in vitro. ^40^ However, other studies and guidelines reported that there is no strong wivermectin in COVID-19.^50, 51^

In the present study we found that steroid use was protective against in-hospital mortality. The findings were the same as the results of review done by WHO Rapid Evidence Appraisal for COVID-19 Therapies (REACT) Working Group. ^38^ Use of steroid is believed to reduce COVID-19 induced inflammatory response and hence halt the pathogenesis of the disease. This in turn reduces lung damage and improve clinical outcomes and prevent mortality.^52^ However, Remdesivir use did not prevent mortality in our study in Tanzania. This could be due to low number (only 9%) of patients who used it in our patients.

Use of supplemental oxygen was associated with mortality in the present study. The use of supplemental oxygen would mean more severe disease and it is well known that correcting hypoxemia sometimes without hypoxia tends to have destructive effects and impairs lung healing. ^53, 54^ Excessive and prolonged oxygen administration can lead to accumulation of reactive oxygen species (ROS) which might lead to progressive destruction of alveolo-capillary membranes in the lungs and cause obstruction of lung capillaries which may forms microthrombi, again due to damage of the alveoli air will lead to the surrounding tissues.^54^ This lung damage by excessive oxygen in combination with COVID-19 can exacerbate cell apoptosis at the alveolar epithelium level resulting in more pulmonary injury and death may occurs^53^.

The study has described clinical manifestations and outcomes of patients diagnosed with COVID-19 infection in Tanzania. However, the limitation of study was its retrospective nature which based on documented patients records, with a large number of patients left out of the analysis due to missed data, this might in a way affect the results of the present study.

## Conclusions

In the present study patients with COVID-19 presented more or less the same as patients elsewhere in the world, with dyspnea, cough, fever, fatigue, chest pain and headache being the most presenting symptoms. Patients who died significantly had higher median value of CRP, D-dimer, white blood cell count (WBC), absolute neutrophil count (ANC), higher fasting blood glucose (FBG), serum creatinine and blood urea nitrogen (BUN). Predictors of mortality were age above 60 years, being uninsured, HIV infection, dyspnea, chest pain, neutrophilia. Steroid use and ivermectin use were found to be protective against COVID-19 mortality. Clinicians should actively look for the predictors of mortality and take appropriate management to reduce mortality.

## Data Availability

No legal or ethical restriction

## Acknowledgement

We acknowledge the Director of the National Public Health Laboratory (NPHL) Mr. Medard Bayega in Tanzania for allowing us to access the COVID-19 database for PCR-results. We would also like to acknowledge Management of Muhimbili National Hospital, Kilimanjaro Christian Medical Center, Bugando Medical Center, Benjamin Mkapa Hospital and Mbeya Zonal Referral Hospital for allowing us to access patients databases. I would also like to acknowledge Evarist Msaki, Rafael Shayo, Amulike Mwakitalu and Anna Jazza for their coordination of data collection.

## Author’s Contribution

Elisha Osati designed the study. Elisha Osati, Naveeda Adams, Dr. Athumani Ramadhani, Mary Nicolaus, Denis Rainer, Christian Mbije, Martha Nkya, did data collection.

Elisha Osati, Grace Shayo, Kajiru Kilonzo, John Meda, Gervas Nyaisonga, Bahati Wanja, Albert Muniko Paulo Mhame, Lucy Samwel, Liggyle Vumilia, Abel Makubi, Tumaini Nagu, Seif Shekalaghe supervised and guided the process of data collection and analysis. Elisha Osati, Raphael Sangeda, Nathan Brand, Candida Moshiro and Grace Shayo did analyze the data.

Elisha Osati and Grace Shayo drafted the manuscript. All authors read and approved the final manuscript for publication.

